# ThinkCancer! The multi-method development of a complex behaviour change intervention to improve the early diagnosis of cancer in primary care

**DOI:** 10.1101/2020.11.20.20235614

**Authors:** Alun Surgey, Stefanie Disbeschl, Ruth Lewis, Julia Hiscock, Sadia Nafees, Rebecca J Law, Jessica L Roberts, Annie Hendry, Zoe Hoare, Nia Goulden, Marian A Stanciu, Andrew Carson-Stevens, Stephanie Smits, Kate Brain, Rhiannon T Edwards, Nefyn Williams, Richard D Neal, Clare Wilkinson

**Author notes:** Correspondence: Dr A Surgey.

## Abstract

**Background:** Relatively poor UK cancer outcomes are blamed upon late diagnosis. Despite most cancer patients presenting to their GP with symptoms, diagnostic delay remains a common theme, with many clinical and non-clinical factors responsible. Early diagnosis is key to improving outcomes and survival. This paper reports the multi-method process to design a complex intervention to improve the timely diagnosis of symptomatic cancer.

**Methods:** A review of reviews, survey, discrete choice experiment, qualitative interviews and focus groups, all informed a realist evidence synthesis. This in turn informed the design of a complex intervention, guided by the Behaviour Change Wheel framework using a multi-step process.

**Results:** Key themes from the realist evidence synthesis included effective safety netting at practitioner and practice system level, increased vigilance and lowering referral thresholds. Qualitative findings explored the tensions, barriers and facilitators affecting suspected cancer referral. The Think Cancer! intervention is an educational and quality improvement workshop directed at the whole primary care team. Bespoke cancer safety netting plans and appointment of cancer champions are key components.

**Conclusions:** Think Cancer! is a novel primary care early cancer diagnosis intervention, requiring evaluation through a cluster randomised control trial.

## Background

The role and importance of primary care (PC) in the diagnosis of cancer is well established.^1^ Wales, like other UK countries, has relatively poor cancer outcomes^2 3^ with late diagnosis recognised as a major contributor resulting in low one-year survival rates.^4 5^ Early diagnosis has been shown to be key in improving cancer outcomes^6^ and cancer survival.^7^ It is therefore unsurprising that national and international cancer policies are heavily focussed on diagnosis. Referral rates and adherence to guidelines are lower in Wales,^8 9^ with General Practitioners (GPs) in the UK overall less likely to take action on potential cancer symptoms.^2^ These findings are unlikely to be unique to Wales with expected similar findings in other countries with comparable health care systems and areas of deprivation.

Most (85%) cancer patients initially present to their GP with symptoms,^10 11^ yet diagnostic delay remains a common and emotive theme in cancer research.^12^ Access to secondary care diagnostics and treatment within the UK health services, as with many other countries with similar models of care, places the GP as the first point of contact in a ‘gatekeeping’ role. Therefore, irrespective of scientific advances such as new diagnostic tests, rapid diagnostic centres for vague symptoms and evolving cancer therapies, unless the GP thinks about the possibility that their patient may have an underlying cancer, delays in investigation and referral will continue.

The Wales Interventions and Cancer Knowledge about Early Diagnosis (WICKED) Programme (Figure 1) aims to develop and test a behavioural intervention targeted at GPs and primary care teams to improve early diagnosis of cancer using the Medical Research Council’s framework for developing and evaluating complex interventions.^13^ The aims of this phase of the project are to synthesise current evidence and to develop a complex intervention that will improve the quality of PC practitioners’ knowledge, attitudes and referral behaviour (work packages 1-3). This paper describes the findings of those work packages and their contribution to intervention development, resulting in the production of a complex behaviour change intervention that will be tested in a feasibility trial (work package 4).

**Figure 1.**
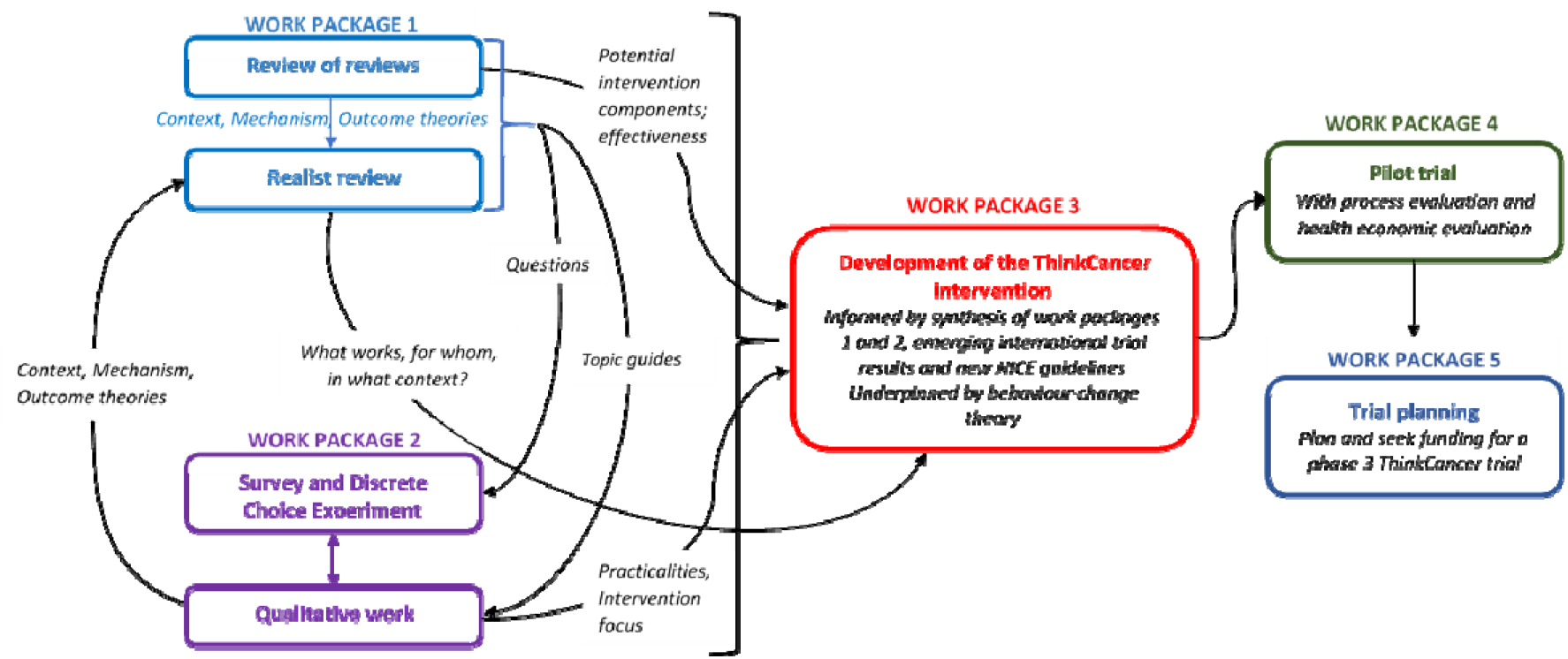
Diagram of the WICKED Programme

## Method

The background, rationale, design and methods of the WICKED programme, leading to the development of the ThinkCancer! intervention, have been reported elsewhere. ^14^Therefore, this section summarises the methods, focusing on their interconnectivity with the intervention development process.

### Work package 1a: Review of reviews

A systematic review of reviews was undertaken to:

- identify the reasons for (or causes of) longer primary care intervals (PCIs) in the diagnosis of cancer;
- identify all interventions designed to reduce the PCI;
- identify the causes of longer PCIs, which could be modified by a behaviour change intervention;
- identify interventions designed to modify the behaviour of PC health professionals;
- identify non-behaviour change intervention components that could contribute to the ThinkCancer! intervention (e.g., structural or organisational modifications that could facilitate the behaviour change elements); and,
- identify which types of interventions (or intervention components) are applicable to UK and similar health systems.

The review of reviews followed methodological guidelines for rigorous conduct and reporting.^16 17^ The search strategy and rationale are reported in the protocol paper,^14^ and are included in detail (supplementary file 1). The searches were conducted in October 2016 and extended to reviews published from 1995 onwards. These searches have not been updated as data was used to inform the starting point of the later intervention development steps, rather than as a stand-alone research output. Two reviewers independently screened titles and abstracts and then the full-text of relevant papers for inclusion. Disagreements were resolved by discussion. Data were extracted in tabular format. We did not formally appraise or exclude studies on the basis of quality; where relevant data contributed to the realist review we considered their ‘trustworthiness’ (i.e., the plausibility and coherence of the methods used to generate the data, and its contribution to the development of the synthesis) in that context.^18^

### Work package 1b: Realist review

The realist review aimed to identify and test programme theories underpinning identified interventions, and determine what works/or not, for whom and in what contexts. This process informed, and was informed by, components of work package 2, and aided the design and development of the ThinkCancer! intervention. Specific objectives were to:

- identify evidence about interventions promoting prompt diagnosis/referral of suspected cancer in PC settings;
- identify the mechanisms through which these interventions directly or indirectly effect changes in the behaviour of individuals, groups and organisations in PC that lead to earlier referral and diagnosis;
- investigate the contextual characteristics that mediate the potential impact of these mechanisms on early referral and diagnosis in PC; and,
- develop a practical programme theory from the evidence synthesis that can be applied to the development of the ThinkCancer! Intervention.

The realist review followed on from the systematic review of reviews, in agreement with Rycroft-Malone, et al., ^19^ adapted from Pawson, et al. ^20^ and RAMESES methodological guidance and reporting standards.^21 22^ The first stage of the process involved data collection from multiple sources organised into a matrix of evidence (supplementary file 2), starting with a list of possible causes of longer PCIs in cancer referral, and examples of interventions to promote prompt diagnosis/referral drawn from scoping searches and background reading. Evidence was mapped against categories of barriers and facilitators to timely referral and diagnosis, further developed through a process of discussion and consensus between the research team operational group (see Figure 2).

**Figure 2.**
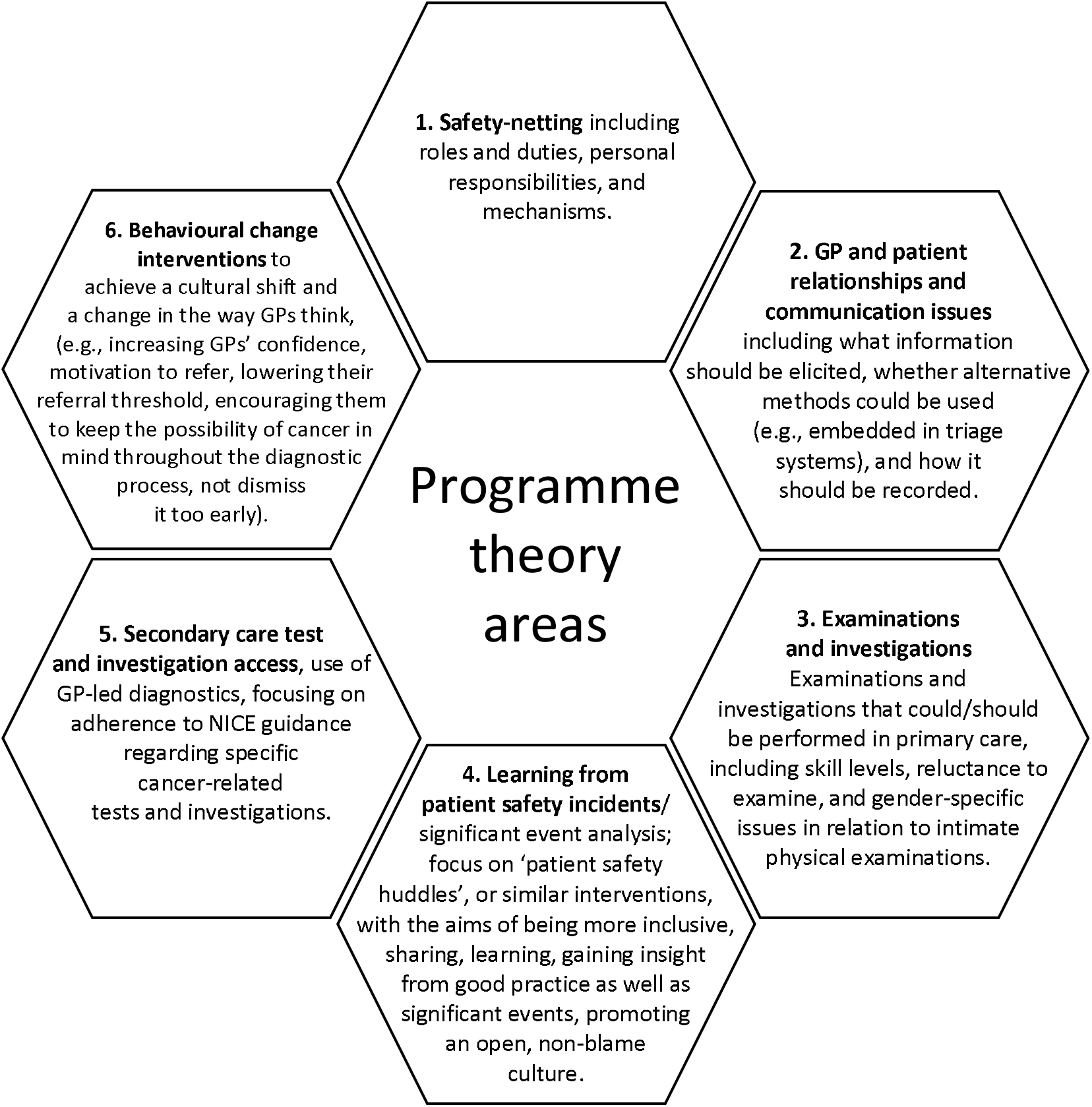
Areas of programme theory used in realist review.

The six areas of focus were then developed as theory areas for the realist review in the form of conjectured Context-Mechanism-Outcomes (CMOs) by two experienced researchers with a background in primary care cancer research, and further refined through several stakeholder group meetings of clinicians and patient participant involvement (PPI) representatives. The final stage of the realist review involved testing and refining the programme theory by searching for and compiling further evidence. A realist narrative was written up for each programme theory area with recommendations.

### Work Package 2a: Quantitative Survey and Discrete Choice Experiment (DCE)

The survey measured the GPs’ cancer knowledge, investigation and referral attitudes, along with their behaviours, learning needs and preferences, and mechanisms supportive of change. The findings were used to identify the influences of Capability, Opportunity and Motivation on the target behaviour (COM-B). COM-B describes how changing behaviour at the individual and/or system level is a result of changing one or more components of psychological and physical Capability, social and physical Opportunity, and automatic and reflective Motivation which informed subsequent intervention development.^14^

All GPs in Wales (approximately 2000) were sent an email invitation to complete the survey between 22^nd^ August 2017 and 18^th^ December 2017. A DCE was part of the online survey. The respondents were required to state preferences regarding hypothetical scenarios,^23^ in order to explore GPs’ preferences for different attributes relating to the earlier diagnosis of cancer in PC, and to explore how GPs are willing to trade between attributes. The attributes and levels were identified from a scoping search, results of which included a review of reviews, experts’ opinions and consensus. Seven attributes used were prompts, audit, type of education and delivery, mode of delivery of education and training, access to diagnostic testing, safety netting and time spent on activities to achieve timely cancer diagnosis.

### Work Package 2b – Qualitative component

The qualitative component explored underlying GP and practice team beliefs and behaviours in identifying, investigating and referring cancer signs and symptoms (including perceived constraints, barriers or concerns), through telephone interviews and focus groups. The purposive sampling procedure and semi-structured topic guide are described elsewhere.^14^ The focus groups also contributed to understanding the practicalities of how the intervention should be focused.

Twenty GPs were interviewed and four focus groups with PC practice teams were conducted. Both data sets were transcribed verbatim, organised and analysed using the five stages of the Framework process: familiarisation, thematic framework identification, indexing, charting, mapping and interpretation.^24^

### Work package 3 – Behaviour Change Wheel (BCW) driven intervention development

All data from work packages 1 and 2 were collated to inform the BCW process and guide the development of an intervention designed to change a target behaviour (see Figure 3). ^14,25^ The starting point in designing a behaviour change intervention is to clarify the target behaviour, or the ‘behavioural diagnosis’.^25^ For ThinkCancer! this was refined according to the main aims of the programme and discussed within the research team and wider stakeholder group. Our proposed target behaviour was identified as *GPs thinking of and acting on clinical presentations* that could be cancer*.

**Figure 3:**
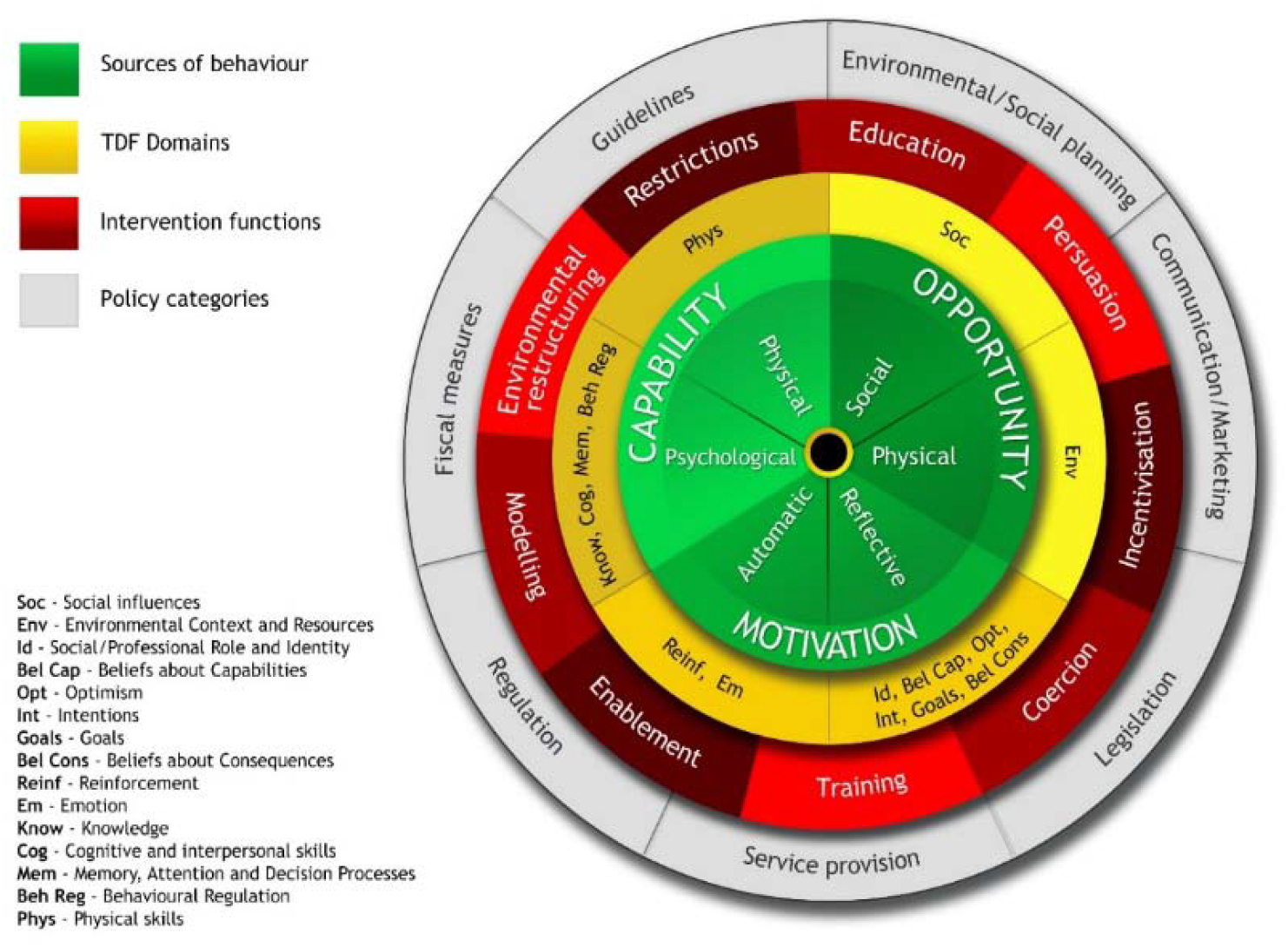
The Behaviour Change Wheel (Reproduced with permission from authors)

Figure 3 depicts the different layers of the BCW with the COM-B model at its centre. (see supplementary file 3 for the detailed methodology of the application of the BCW).

Using a matrix of intervention functions and applying the APEASE criteria (Affordability, Practicability, Effectiveness/cost-effectiveness, Acceptability, Side effects/safety and Equity) ^25 26^ we selected the following functions that best suited our target behaviour: education, training, enablement, environmental restructuring, modelling, persuasion and incentivisation. Intervention functions were then matched to relevant Behaviour Change Techniques (BCTs), for example, in relation to education and training, the relevant intervention functions included: *instruction, information about health consequences, feedback on outcomes, prompts/cues, self-monitoring, and verbal persuasion about capability*. A multidisciplinary intervention working group was convened to develop the intervention, including members of the research team, a wider stakeholder group including GPs to ensure real world applicability and an expert patient.

### Public and Patient Involvement (PPI)

There has been continued and active Public and Patient Involvement (PPI) within the research programme from its initial design and continued through each work package, with regular contribution to research group meetings and advice and comment gained on project documentation.

## Results and Intervention Development

### Work package 1a: Review of reviews

After discarding duplicate references, 334 studies were identified, of which 37 were deemed to be potentially relevant and full papers were retrieved. Seventeen of these were included in the review. ^7 27-42^ A summary of findings is presented in supplementary file 4.

Thirteen of the included studies were systematic reviews ^27 29 31-36 38-42^ and three were non-systematic narrative literature reviews.^7 30 37^ The final study reported a consensus process with key informants that was based upon an earlier literature review.^28^ Ten studies focused on specific cancers: haematological, breast and colon;^27^ bladder ^29 39^; colorectal ^30 33 41 42^; upper gastro-intestinal ^31 32^; and colorectal and lung. ^36^ The remaining seven considered any cancer. Broadly speaking, three studies focused on the impact on cancer referral of features of healthcare systems, ^28 33 37^ two focussed on clinical roles and practices, ^29 31^ two on signs, symptoms and the process of diagnosis, ^39 40^ and six on identifying or describing factors relating to diagnostic intervals.^7 27 30 32 35 36^

Only three reviews aimed to include studies that evaluated interventions to address delays or promote early diagnosis of cancer.^32 34 38^ Macdonald, et al.^32^ evaluated factors associated with longer patient and PC practitioner intervals for upper gastrointestinal cancer and aimed to describe an intervention designed to reduce those intervals, but found no intervention studies that met their inclusion criteria.^32^ Mansell, et al.^34^ aimed to identify interventions that reduce PC delay. Although the review covered any cancer, only skin, breast, colorectal studies met the inclusion criteria; 22 interventions were identified, but no evidence that any intervention directly reduced PC delay in the diagnosis of cancer. Limited evidence suggested that complex interventions such as audit and feedback, and specific skills training had the potential to reduce delays. Schichtel, et al.^38^ examined the evidence for the effectiveness of educational interventions for primary healthcare professionals. Twenty studies were included but the authors concluded that, although some educational interventions delivered at a clinician as well as at a practice level may promote the early diagnosis of cancer in PC, there is limited evidence for their long-term sustainability and effectiveness.

### Work package 1b: Realist review

The following initial conjectured Context-Mechanism-Outcome configurations (CMOs) were developed and refined:

CMO 1: Where explicit systems are put in place to plug potential gaps in the patient pathway (i.e. safety netting) **(context)**, and members of the healthcare team identify and perceive themselves to have safety-netting responsibilities that are important and relevant to their role (e.g. to ensure test results are acted upon, referrals sent, follow-up appointments made and attended as advised etc.) **(mechanism)**, then they will act upon them, and patients will be more likely to progress to referral without <u>undue</u> delay **(outcome)**.

CMO2: Where systems are in place and patients are engaged in safety-netting processes (e.g. by being advised to come back if symptoms are not resolved or get worse, asked to ring if they don’t hear about a test result, hospital appointment etc.) **(context)**, and, by becoming more involved, they perceive the importance of these actions and are empowered to take more responsibility **(mechanism)**, then they will adopt a more proactive role in their care and will be less likely to miss appointments etc. **(outcome)**.

CMO 3: Where all practice staff who have contact with patients have been trained to recognise unexpected signs and uncharacteristic behaviours (e.g. altered appearance such as weight-loss, unusual consulting pattern) **(context)**, and are therefore alert to, and notice these signs, and recognise the possibility of cancer **(mechanism)**, then staff will be more likely to question patients and report suspicions, and GPs may identify more patients who they *should* worry about, pay closer attention to these patients, and perhaps elicit important information (signs, symptoms, previous or family history etc. that the patient may not recognise as relevant and would not otherwise volunteer) **(outcome)**.

CMO 4: Where usual practice includes discussion of patient-safety (e.g. ‘safety huddles’, significant event audits), and there is a culture of frank, informal, non-judgemental interaction **(context)**, and by sharing experiences and supporting each other, GPs benefit in terms of improved knowledge and confidence in their actions **(mechanism)** then they will be more likely to recognise when investigations and referrals are warranted, and less likely to delay referral because of uncertainty **(outcome)**.

CMO 5: Where a practice audit of cancer referrals is conducted and feedback is given to GPs on whether recommended investigations and examinations were conducted **(context)**, they will be aware of and reflect on their own personal performance, compared with local or national performance, and recognise how it can be improved and **(mechanism)** then they will be more likely to perform these tasks consistently, in line with good clinical practice and NICE guidance **(outcome)**.

CMO 6: In the face of uncertainty about individual cancer referral decisions (e.g. vague symptoms, low-risk but not no-risk), if a philosophy of cancer-risk-averseness, and a practice culture of case-finding rather than gate-keeping are engendered **(context)**, GPs will be more likely to lower their individual risk threshold for urgent referrals and Think Cancer! **(mechanism)**, so that more cases of suspected cancer will be identified and referred **(outcome)**.

The final stage of the realist evidence synthesis led to completion of narrative summaries (Supplementary File 2), summarised into the following key headlines:

- Wider use of high quality and effective safety-netting in every aspect of the patient/health care professional interaction;
- Dual safety-netting responsibility between patient and practitioner by encouraging aware, engaged and empowered patients to co-navigate the patient journey;
- Increased vigilance and taking action when suspicions are raised (including all practice staff);
- Improving the practice level patient safety agenda;
- The right test, for the right patient, at the right time (guidelines, awareness, false negatives); and,
- Lowering the threshold to referral trigger (challenging gate keeper mentality, acting on gut feelings).

### Work package 2a: Quantitative Survey

In total, 1993 GPs were invited and 269 responded (13.5%). Respondents were representative of GPs in Wales in terms of personal and practice factors. There was strong agreement amongst respondents that they had changed their behaviour with regard to safety netting of symptoms in the past few years, and changing thresholds for investigation and referral, but less so for the use of decision support tools. The most prominent drivers of change were case discussions with colleagues and significant event audits. There was strong agreement that timelier diagnosis leads to better survival for the six cancers included in the survey (breast, colorectal, endometrial, myeloma, pancreas and renal), and that GPs had a large amount of influence on timely diagnosis of these cancers. Respondents were less confident about patient management when they did not have qualifying symptoms for urgent referral. GPs conveyed that they can, and have, changed their cancer-related diagnostic activity, and believe that they can influence timely diagnosis. They also identified a number of ways in which future interventions may be targeted to maximise behaviour change.

**Table 1.** Exploring Capability, Opportunity and Motivation for behaviour change COM-B

| **When it comes to you personally referring patients with suspected cancer symptoms, what do you think it would take for you to do it even better** |  |  |  |  |
| --- | --- | --- | --- | --- |
| I would have to.... | N | Missing | Yes | No |
| Capability |  |  |  |  |
| Know more about why it was important | 253 | 16 | 85 (33.6%) | **168 (66.4 %)** |
| Know more about how to do it | 253 | 16 | 71 (28.1%) | **182 (71.9%)** |
| Have better physical skills | 253 | 16 | 60 (23.7%) | **193 (76.3%)** |
| Have more mental strength | 253 | 16 | 67 (26.5%) | **186 (73.5%)** |
| Overcome mental obstacles | 253 | 16 | 94 (37.2%) | **159 (62.8%)** |
| Have more mental stamina | 253 | 16 | 101 (39.9%) | **152 (60.1%)** |
| Opportunity |  |  |  |  |
| Have more time to do it | 253 | 16 | **195 (77.1%)** | 58 (22.9%) |
| Have more money | 253 | 16 | 19 (7.5%) | **234 (92.5%)** |
| Have it more easily accessible | 253 | 16 | **218 (86.2%)** | 35 (13.8%) |
| Have more people around me doing it | 253 | 16 | 90 (35.6%) | **163 (64.4%)** |
| Have more triggers to prompt me | 253 | 16 | 110 (43.5%) | **143 (56.5%)** |
| Have more support from others | 253 | 16 | **154 (60.9%)** | 99 (39.1%) |

| Motivation |  |  |  |  |
| --- | --- | --- | --- | --- |
| Feel that I want to do it enough | 252 | 17 | 55 (21.8%) | **197 (78.2%)** |
| Feel that I need to do it enough | 252 | 17 | 83 (32.9%) | **169 (67.1%)** |
| Believe that it would be a good thing to do | 252 | 17 | 126 (50.0%) | 126 (50.0%) |
| Develop better plans for doing it | 252 | 17 | **142 (56.3%)** | 110 (43.7%) |
| Develop a habit of doing it | 252 | 17 | **139 (55.2%)** | 113 (44.8%) |

\*Bold entries indicate the highest response category

### Work package 2a: Discrete Choice Experiment (DCE)

Of the 269 survey respondents, 151 fully completed the DCE. Seven attributes and preferences were explored, with five showing statistical significance. Open access to diagnostic tests was ranked as most important, followed by practice level cancer audit. The other statistically significant preferences were computerised prompts, blended online and face to face education sessions and a preference towards patient responsibility in safety netting. The focus of educational continuing professional development and time spent doing it were not ranked as statistically significant attributes. (Supplementary File 5 shows the attributes and attribute levels offered along the logit regression model and detailed DCE data analysis).

### Work package 2b: Qualitative component

The qualitative work undertaken as part of the WICKED programme revealed important insights and rich data relating to current challenges of PC teams dealing with patients who might have cancer. A separate scientific paper is proposed to illustrate the methods and findings in detail. To highlight some common themes with regard to intervention development, participants explained how they go about identifying, investigating and referring patients with potential signs and symptoms of cancer.

They described how they address the need to diagnose cancer earlier, and what influences their behaviour, including NICE guidance, clinical judgement, patient factors and ‘gut feeling’.

“*Well, some of them [the guidelines] you use every day, so it just becomes*…*something you’re thinking about constantly*…*just part of the everyday decision making*.” (Male, urban, medium deprivation, < 10yrs experience)

However, they were also influenced by their knowledge of the pressures within secondary care, and a concern not to add to an already strained system. This created a tension between GPs’ internal, individual decision-making and their knowledge of the context-dependent pressures.

“*A big part of me wants to refer everyone. Yeah, 3% risk, you’re more likely to catch that earlier diagnosis and get a curable disease. But at the same time, you don’t want to completely swamp the system so that no one’s getting it. It is very resource-limited. It does affect my practice significantly*.”

(Female, urban, high deprivation,15-25yrs experience)

Exploring perspectives from practice teams during the focus groups, it appears that some of this tension can potentially be offset by positive practice culture and helpful practice-based systems. For example, open door communication systems between staff members and a feeling that the whole practice team is united in a common goal.

“*I think on the phone you don’t see people do you, but at the desk you do. So you notice their weight, or the yellowness or, you know if you think there is something not right, I mention it to one of the doctors*.”

(Receptionist, urban area, non-training practice)

### The novel intervention -ThinkCancer!

Data from all earlier work packages were included to build the content and proposed modes of delivery for a host of potential interventions. The recurring themes of safety netting, use of guidance and lowering referral thresholds were strongly represented with regard to content of the intervention, among other key topics. The most common intervention functions identified through the BCW process were education, training, enablement and environmental restructuring.

The resultant ThinkCancer! Intervention (see Figure 4) is a whole practice-based educational and quality improvement workshop, consisting of themed sessions for both clinical and non-clinical staff, the co-production of a bespoke practice-specific Cancer Safety Netting Plan (CSNP) and the appointment of a Cancer Safety Netting Champion (CSNC). The workshop aims to raise awareness and increase knowledge around current cancer diagnosis guidance and be delivered over half a day during GP protected educational time in the form of face-to-face or remotely delivered web based educational sessions. Sessions will consist of a series of interactive activities exploring existing processes within practices and developing plans for change to implement each component. Members of the research team will deliver the intervention; a member qualified as a GP Educator will oversee the workshop, supported by up to two research team staff members.

**Figure 4.**
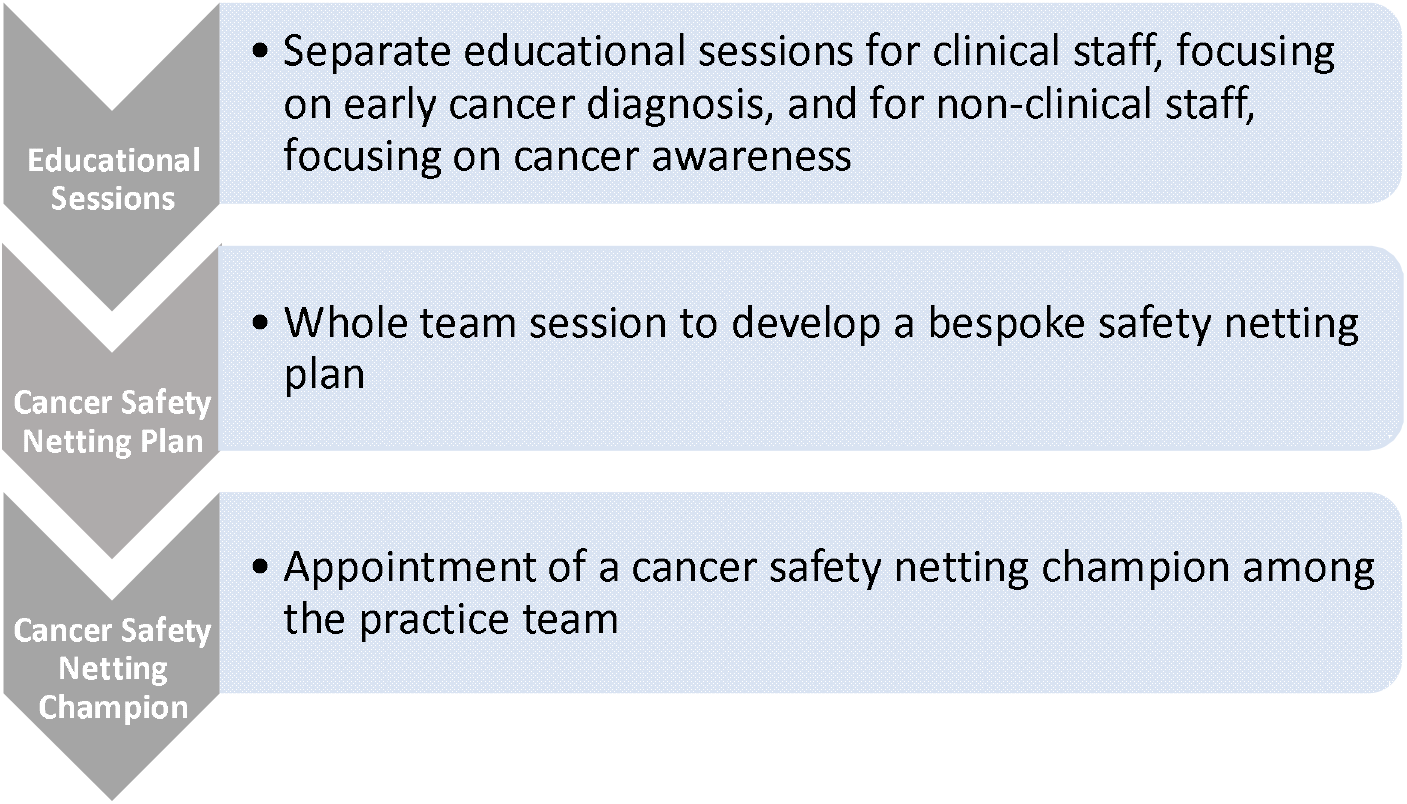
Summary outline of the ThinkCancer! intervention

#### Session 1

This session of the workshop will deliver a face-to-face or remote video linked educational session for all clinical staff at the practice. The overall aim is to raise knowledge and awareness around current guidance, hot topics in early diagnosis and safety netting, including the introduction of a new safety netting tool – the Shared Safety Netting Action Plan (SSNAP). The educational component of the ThinkCancer! Intervention has been developed in accordance with well-established educational principles and theories such as social cognitive theory and andragogical assumptions. ^43^The curriculum principle, constructive alignment, was considered in mapping the intended learning outcomes (ILOs) against the learning activities and experiences, along with the assessment and evaluation plan. ^44^ Opportunities for discussion and reflection within the workshops will allow learners to connect any new learning with prior experience, ^45^and making clear the relevance of the learning programme to daily practice should increase both the engagement and effectiveness of the learning experience. The session will specifically focus around key Intended Learning Outcomes (ILOs) as described below. The ILOs were developed from stakeholder review of data collected from work packages 1 and 2, specifically the programme theory developed from the realist review. This was later further developed and refined in collaboration with RedWhale GP Update™ who are an established organisation delivering PC update courses, including a specific not for profit cancer and palliative care training programme. Hot topics in early diagnosis of cancer and common pitfalls were included in the ILOs from their materials. This collaboration allowed educational and content oversight and peer review. The ILOs for the clinician session comprise the following:

1. Understand the key points about the latest NICE guidelines;
2. Consider the recurrent themes in delayed diagnosis;
3. Be aware of guidance around thrombocytosis and cancer;
4. Be alert to common pitfalls in cancer detection and the dangers of false negative investigations;
5. Understand the importance of and techniques considered essential for high quality safety netting, with introduction of the safety-netting prescription;
6. Use the *ThinkCancer!* toolkit/manual as an information and signposting resource.

##### Shared Safety Netting Action Plan (SSNAP) Tool

The SSNAP tool is a co-designed novel safety netting intervention in the form of a physical leaflet that can be given to a patient that formalises the advice given within a consultation detailing when and how to seek further medical review. Developed by a research team within the Yorkshire and Humber Patient Safety Translational Research Centre, the tool was developed from a mixed-method programme of research involving systematic review and exploratory qualitative work,^46 47^ and culminating in a series of iterative co-design workshops and focus groups, including PC teams and patients. The use of the tool within the ThinkCancer! intervention practices and incorporation into their own bespoke safety netting plan is at the discretion of the individual practice.

#### Session 2

The non-clinical session will focus on patient-facing staff and their role in the early diagnosis of cancer. The aim is to increase awareness of red flag symptoms (using the Cancer Research UK Be Clear on Cancer Campaign)^48^ and highlight non-specific but concerning symptoms that patients may complain of or present, that could indicate early presentation of cancer.^49^ The secondary aim of this workshop is to improve confidence in sharing concerns within the practice team by placing the patient at the centre of all healthcare interactions and empowering staff. The format will be interactive and in two parts. Part 1 will include a gallery of posters on the wall with a walk around and discussion focussed on the Be Clear on Cancer campaigns. Symptom cards will be given that will include red flag symptoms for possible cancer and also other common symptoms reception staff are faced with when interacting with patients. Participants will then be asked to separate the symptom cards into two groups; red flag versus non red flag for cancer symptoms. The format for the matching exercise was conceived by a member of the research team with experience in adult and medical education. The second part will be a case discussion exploring common themes in early presentation of cancer, such as patient behaviours, for example changes in consultation patterns.

#### Session 3 – Cancer Safety Netting Plan (CSNP) and Cancer Safety Netting Champion (CSNC)

The aim of this component of the workshop is to facilitate the design and implementation of a practice-specific Cancer Safety Netting Plan (CSNP). This component of the workshop will be attended by both clinical and administrative staff. Information gathered prior to the workshop from a baseline practice questionnaire regarding current safety netting procedures will be discussed and reflected upon in an exercise involving the use of the CRUK safety netting flow diagram (Supplementary File 6). The practice team will consider the practice’s existing safety netting arrangements (if any), and the need for modification, augmentation, or the development of an entirely new plan. Overall leadership and responsibility for implementing the plan will rest with the practice-appointed Cancer Safety Netting Champion (CSNC), who will ideally be identified before the workshop but may be identified during the workshop. The use of a champion to affect positive change in projects and organisations is now well established.^50^ The CSNC will take ‘ownership’ of the CSNP and be responsible for implementing it within and throughout the practice as well as the encouragement of a supportive environment for staff. The production, implementation, monitoring and evaluation of the plan will follow recognised Quality Improvement (QI) principles.^51 52^ However, the minimum output of the intervention will be the reflective discussion generated and testing whether the development of a safety netting plan is feasible.

#### Logic model

In order to assess and evaluate ThinkCancer!, drawing on the UK MRC framework for evaluation of complex interventions,^13^ a logic model of the proposed intervention has been created (Figure 5). This will allow the research team to gain a greater understanding of each working part of the intervention by mapping out each individual component of the intervention by its input and activity. Detailing the anticipated outputs and outcomes allow the research team to consider the most effective ways of measuring and recording the effects of the intervention, therefore suggesting primary and secondary outcome measures as featured in this logic model. The current primary outcome measures for ThinkCancer! are Two Week Wait referral rate and Primary Care Interval (PCI), where the assumption is made that raising awareness and providing education will raise referral rates, and improving safety netting and GP practice systems will shorten the PCI, thereby reducing diagnostic delay in the more difficult to diagnose cancers. These measures are expected to best assess effectiveness of the intervention during a definitive phase 3 randomised controlled trial. The secondary outcome measures relate to feasibility criteria as this is a novel complex intervention.

**Figure 5.**
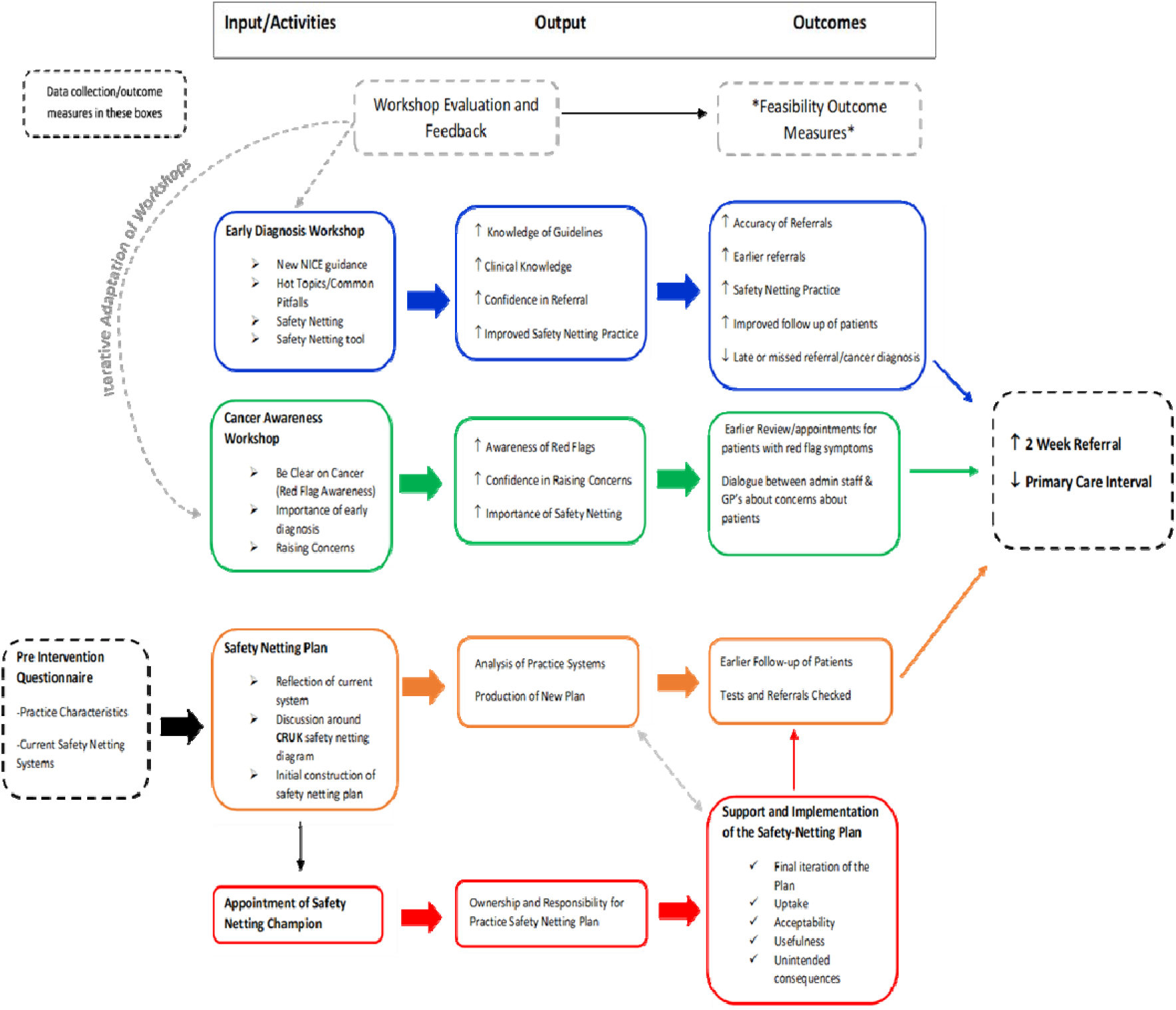
The ThinkCancer! Intervention Logic Model

Refinement of the logic model from data collected during a proposed feasibility trial will allow iterative development of the intervention.

## Discussion

The WICKED programme involved a rigorous and structured process of intervention development, built on a sound foundation of primary research, by a core team of multi-professional researchers supported by methodological specialists. The process was guided at each stage by expert patient involvement and stakeholders, contributing to the validity and future implementation of the intervention. This research programme was originally set up to improve cancer outcomes in Wales, and has developed an intervention that is relevant across the whole of the United Kingdom and for any other country where health care is structured around primary care as the first point of contact. The landscape of cancer diagnosis is topical and fast changing with research and policy change occuring at a rapid rate. Single pathway referral processes may aid referrers and patients along their diagnostic journies when cancer is suspected, and the introduction of Rapid Diagnostic Centres for pateints prssenting with low risk but not no risk symtpoms are likely to reduced diagnostic intervals. However, if the possibility of a cancer diagnosis is not considered, if the clinician does not Think Cancer!, the patient will not be referred and diagnostic delays continue.

This potential limitation to the validity of the data collected in the early review work has been mitigated by the research team regularly reviewing new published papers and policy against the original findings. The strength of this multidimensional approach applied in the WICKED programme lies in two important factors. Firstly, the deep insight gained within the qualitative work in revealing the attitudes, feelings and behaviours of the clinical staff in relation to real world challenges of cancer diagosis and referral, giving sharp focus to what is most important within the literature to exploit in an intervention to promote timely cancer diagnosis. Secondly, in the use of the BCW to guide the intervention development, all data from each work package was recycled to inform the process, eliminating any “research waste”. This environmentally friendly research method has added to the strength, depth and scientific rigour of the format and delivery of the proposed intervetion. It is difficult to objectively measure the quality of intervention development and reporting but there are limited guidelines, such as The Template for Intervention Description and Replication (TIDiER) checklist.^53^ ThinkCancer! satisfies all domains relating to initial design prior to testing as described in this check list. A recently published eDelphi consensus aiming to provide more indepth guidance on the reporting of intervention development studies in health research (GUIDED),^54^ achieved consensus on fourteen reporting items. The intervention development described in this paper satisfies the majority of those reporting items. ThinkCancer! will be tested and iteratively developed within a feasibility trial delivered in Wales in 2020, with an economic and mixed method process evaluation.

## Conclusion

ThinkCancer! is a novel, theory-informed educational and quality improvement practice-level intervention, aiming to improve the early diagnosis of cancer in primary care. The content of the intervention and the way it will be delivered has been the output of a comprehensive multi-method programme of primary research, developed in accordance with the MRC framework.^13^ This complex intervention will be tested in a randomised controlled feasibility trial, with embedded process and economic evaluation, to iteratively adapt and refine the intervention, assess the usual feasibility criteria and test proposed outcome measures. The findings will go on to inform a definitive large-scale phase 3 randomised control trial.

## Data Availability

Supplementary files referred to in this manuscript are attached.

## Additional Information

### Authorship

All named authors meet the International Committee of Medical Journal Editors (ICMJE) criteria for authorship for this article, take responsibility for the integrity of the work as a whole, and have given their approval for this version to be published.

### Authors’ contribution

The authors contributions are as follows: conceiving and designing the study: CW,RDN,NW,RTE, KB, SS, ACS, MAS, NG, ZH, RJL, SN, JH, RL; data gathering and interpretation: CW,RDN, NW, RTE, KB, SS, ACS, MAS, NG, ZH, AH, JLR, RJL, SN, JH, RL, SD, AS; drafting or revising the manuscript CW, RDN, NW, RTE, KB, SS, ACS, MAS, NG, ZH, AH, JLR, RJL, SN, JH, RL, SD, AS.

### Data availability

All data generated or analyzed during this study are included in this published article/as supplementary information files.

## Acknowledgements

The authors would like to thank our patient representative Jan Rose and all clinical and research colleagues who assisted in the design, delivery and data interpretation of the WICKED research programme. We acknowledge the contributions of Maggie Hendry (programme design and review work), Dr Lynne Williams (realist review), Dr Jane Heyhoe (Yorkshire and Humber Patient Safety Translational Research Centre -intervention development and process evaluation) and Dr Caroline Greene (RedWhale GPUpdate – intervention development and medical education expertise).

