## Supplementary File 1 for "ThinkCancer! The multi-method development of a complex behaviour change intervention to improve the early diagnosis of cancer in primary care"

Supplementary File 1 – Review of reviews search strategy

*Search strategy developed for Medline and adapted for the remaining databases*

1. Exp neoplasms/
2. (Neoplasm* or cancer* or carcinoma* or malingnan* or tumo?r).ti,ab,kf,kw.
3. 1 or 2
4. (Delay* or time or interval) adj4 (diagnos?s or referral*).ti,ab,kf,kw.
5. (Early or timely or fast* or rapid or prompt or expedite) adj4 (diagnos?s or referral*).ti,ab,kf,kw.
6. 4 or 5
7. Exp family practice/ or Exp primary health care/ or Exp community health services/
8. (General practi* or GP or (primary adj2 care) or (family adj2 physician) or (family adj2 practi*) or (family adj2 doctor) or (community adj2 (service or care)) or (general adj2 physician)).ti,ab,kf,kw.
9. 7 or 8
10. meta-analysis.pt.
11. meta-analysis/ or systematic review/ or meta-analysis as topic/ or "meta analysis (topic)"/ or "systematic review (topic)"/ or exp technology assessment, biomedical/
12. ((systematic* adj3 (review* or overview*)) or (methodologic* adj3 (review* or overview*))).ti,ab,kf,kw.
13. ((quantitative adj3 (review* or overview* or synthes*)) or (research adj3 (integrati* or overview*))).ti,ab,kf,kw.
14. ((integrative adj3 (review* or overview*)) or (collaborative adj3 (review* or overview*)) or (pool* adj3 analy*)).ti,ab,kf,kw.
15. (data synthes* or data extraction* or data abstraction*).ti,ab,kf,kw.
16. (handsearch* or hand search*).ti,ab,kf,kw.
17. (mantel haenszel or peto or der simonian or dersimonian or fixed effect* or latin square*).ti,ab,kf,kw.
18. (met analy* or metanaly* or technology assessment* or HTA or HTAs or technology overview* or technology appraisal*).ti,ab,kf,kw.
19. (meta regression* or metaregression*).ti,ab,kf,kw.
20. (meta-analy* or metaanaly* or systematic review* or biomedical technology assessment* or bio-medical technology assessment*).mp,hw.
21. (medline or cochrane or pubmed or medlars or embase or cinahl).ti,ab,hw.
22. (cochrane or (health adj2 technology assessment) or evidence report).jw.
23. (meta analysis or review).pt.
24. (comparative adj3 (efficacy or effectiveness)).ti,ab,kf,kw.
25. (outcomes research or relative effectiveness).ti,ab,kf,kw.
26. ((indirect or indirect treatment or mixed-treatment) adj comparison*).ti,ab,kf,kw
27. OR/10-26
28. 3 AND 9 AND 27

Note: As the reasons for longer Primary Care Intervals (PCIs) and the possible mechanisms to expedite referral are so numerous, we did not include search terms for specific interventions so that none could be unintentionally omitted. Since an important aim of the review of reviews was to be a starting point of the realist review, and in realist terms relevance is considered to be of paramount importance, we considered any type of review article (i.e., traditional literature reviews as well as systematic reviews).
