## Supplementary File 2 for "ThinkCancer! The multi-method development of a complex behaviour change intervention to improve the early diagnosis of cancer in primary care"

Supplementary File 2 – Realist review final stage evidence review and CMO refinement

Most of the CMOs related to some aspects of safety-netting, we began with literature searches in this area. The following search strategy, developed for Medline and subsequently adapted for CINAHL, PsycINFO, and the Cochrane Library, was run in November 2017:

1. safety netting.mp.
2. case manage$.mp. or Case Management/
3. patient navigat$.mp. or Patient Navigation/
4. primary care.mp. or Primary Health Care/
5. cancer.mp. or Neoplasms/
6. 2 and 4 and 5
7. 3 and 4 and 5
8. 1 or 6 or 7

The searches achieved 147 ‘hits’ after de-duplication. The titles and abstracts were independently screened for relevance, and full texts for inclusion by 2 reviewers. Fourteen were agreed to be potentially relevant, two of which were excluded on review of the full texts; three more were identified by the study team and added. Two reviewers independently extracted study details and data relevant to the CMOs using a bespoke data-extraction form (supplementary file); the two versions were checked for consistency and consolidated. The study details were summarised in tabular format (supplementary file), including comments on the trustworthiness of the evidence, which, overall, was considered very good. Data relating to each CMO were then aggregated into an evidence table, further supplemented by data collected as part of work package two, i.e. the charted qualitative data, and the content analysis of free-text survey responses. The lengthy evidence table was then condensed in a summary version (supplementary files).

Additional papers subsequently identified by the study team, including all papers from a special edition of BJGP focusing on early cancer diagnosis (May 2018), as well as a large number of titles identified during the data-extraction process from the reference lists of included papers, were screened and, if relevant, read. A reference list of the papers read is available in supplementary file xx, but they were not data-extracted as, although relevant, they did not add anything new (i.e. data-saturation had been achieved).

The summarised evidence was further condensed and scrutinised alongside the conjectured CMO configurations. Refined versions of the CMOs were developed, in light of the evidence, initially by two members of the research team, and further refined by an operational group, three of whose members were GPs, through a process of discussion and consensus. The following data tables describe the CMO refinement.

| **Conjectured CMO 1** | **Evidence** | **Refined CMO 1** |
| --- | --- | --- |
| Where explicit systems are put in place to plug potential gaps in the patient pathway (i.e. safety-netting) **(context)**, and members of the healthcare team identify and perceive themselves to have safety-netting responsibilities that are important and relevant to their role (e.g. to ensure test results are acted upon, referrals sent, follow-up appointments made and attended as advised etc.) **(mechanism)**, then they will act upon them, and patients will be more likely to progress to referral without undue delay **(outcome)**. | Evidence suggests that safety netting should include access to and continuity of care*, communication with patients about diagnostic uncertainty*, what signs/symptoms to look for, when and how to seek further help, planned follow-up*, systems for prompt review and action on test results, referrals etc., and consistent documentation* of plans/actions.^1-8^ **(context)** Safety-netting happens when there is a culture of vigilant care*, supported by good communication between practice staff* and with out-of-hours services and secondary care.^2 3^ ^4^ **(mechanism)**  Evidence supports safety-netting as a method of managing diagnostic uncertainty and keeping a track of patients.^1 2 4 5 7-8^ **(outcome)**  Factors that may facilitate/support safety-netting include a standardised process,^3^  training,^3^ good, printable patient information,^3^ a lower threshold for asking patients to return for review,^6^ and greater awareness of hidden symptoms (i.e. masked by co-morbidity or false-negative test results, or hidden patient agendas that emerge late in a consultation).^5 6^  Potential barriers are restriction of access to doctors by reception staff,^2^ and lack of GPs’ time.^3^ Electronic reminders and decision tools are found helpful by some GPs, but annoying by others.* Continuity of care may help safety-netting but sometimes a ‘fresh pair of eyes’ is needed.* | Where explicit systems are put in place to plug potential gaps in the patient pathway (i.e. safety-netting) **(context)**, and there is a culture of vigilant care, supported by good communication between practice staff and secondary care, where members of the healthcare team identify and perceive themselves to have safety-netting responsibilities that are important and relevant to their role **(mechanism)**, then they will use safety-netting as a method of managing diagnostic uncertainty and keeping a track of patients, and patients will be more likely to progress to referral without undue delay **(outcome)**. |
| **Conjectured CMO 2** | **Evidence** | **Refined CMO 2** |
| Where systems are in place and patients are engaged in safety-netting processes (e.g. by being advised to come back if symptoms are not resolved or get worse, asked to ring if they don’t hear about a test result, hospital appointment etc.) **(context)**, and, by becoming more involved, they perceive the importance of these actions and are empowered to take more responsibility **(mechanism)**, then they will adopt a more proactive role in their care and will be less likely to miss appointments etc. **(outcome).** | Evidence suggests that diagnostic uncertainty should be explicitly shared with patients*, and that the GP should ensure it is heard and understood, that patients know what to do/look out for (regarding follow-up appointments, test results etc.), and are involved in discussion/decision-making*.^1 5-8^ **(context)** Patients’ involvement in safety-netting is influenced by the clarity of advice*, level of reassurance* (i.e. not to cause undue anxiety, nor to over-reassure), and the balance of responsibility between patient and GP*,^1 5 9^ but it can be a means of empowering the patient* **(mechanism)**, so that they can take responsibility, monitor their own situation and take effective action when needed.^1^ **(outcome)**  Patient engagement may be influenced on the part of the doctor by their consulting skills/style,^2 3 7^ or their observations (e.g. changes in appearance or behaviour)^2^ or perceptions (e.g. sensible, worrier)^2 9^* of the patient. Patients may alert the GP to the possibility of cancer by mentioning their own suspicions*, or they may play down symptoms so as not to ‘bother’ the doctor.^5 8 10^* It is unclear whether advice should be written or verbal.^3 7^^11^ | Where systems are in place and patients are engaged in safety-netting processes by being fully informed, given clear advice, and engaged in discussion and decision-making **(context)**, and, by becoming more involved, they are empowered to take more responsibility **(mechanism)**, then they will adopt a more proactive role in their care and be more likely to monitor their own situation and take effective action when needed **(outcome).** |
| **Conjectured CMO 3** | **Evidence** | **Refined CMO 3** |
| Where all practice staff who have contact with patients have been trained to recognise unexpected signs and uncharacteristic behaviours (e.g. altered appearance such as weight-loss, unusual consulting pattern) **(context),** and are therefore alert to, and notice these signs, and recognise the possibility of cancer **(mechanism)**, then staff will be more likely to question patients and report suspicions, and GPs may identify more patients who they *should* worry about, pay closer attention to these patients, and perhaps elicit important information (signs, symptoms, previous or family history etc. that the patient may not recognise as relevant and would not otherwise volunteer) **(outcome)**. | Evidence suggests that subtle signs are easily missed and may be normalised/under-reported by patients.^11^* However, altered physical appearance or behaviour can be a trigger for concern,^10^ and all practice staff could be involved in looking for and flagging up suspicious signs to the GP.^2 10^* A chain of communication between all practice staff facilitates this, as different staff members will be familiar with different patients/patient groups.^2 10 11^* **(context)** Practice staff may be more likely to be involved in cancer detection and awareness if they perceive it to be their role.^10^* **(mechanism)** Non-clinical as well as clinical staff may notice and report subtle signs such as altered physical appearance, behaviour, or consulting patterns, and persuade reluctant patients to see a GP if appropriate. They may also take opportunities to educate the practice population about symptoms that should be discussed with a GP.^2 4 5 10-12^ **(outcome)**  Other clinical and non-clinical staff may have the advantage of knowing the patient well (compared with a GP who is new/ a trainee/a locum). The involvement of non-clinical staff in safety-netting may be supported by training e.g. navigation training, red-flag training, and by a non-hierarchical practice structure.* | Where all practice staff who have contact with patients are able to recognise subtle signs and uncharacteristic behaviours (e.g. weight-loss, unusual consulting pattern), which may be normalised or under-reported by patients **(context),** and are therefore alert to, and notice these signs, and recognise the possibility of cancer **(mechanism)**, then staff will be more likely to question patients and report suspicions, and GPs may identify more patients who they *should* worry about, pay closer attention to these patients, and perhaps elicit important information (signs, symptoms, previous or family history etc. that the patient may not recognise as relevant and would not otherwise volunteer) **(outcome)**. |
| **Conjectured CMO 4** | **Evidence** | **Refined CMO 4** |
| Where usual practice includes discussion of patient-safety (e.g. safety ‘huddles’), and there is a culture of frank, informal, non-judgemental interaction **(context)**, and by sharing experiences and supporting each other, GPs benefit in terms of improved knowledge and confidence in their actions **(mechanism)** then they will be more likely to recognise when investigations and referrals are warranted, and less likely to delay referral because of uncertainty **(outcome)**. | Evidence supports the usefulness of sharing knowledge, challenging opinions, discussion, reflection, advice and mutual support. ^5 10 12^* **(context)** The success of safety-huddles (or similar activities) might be influenced by a non-hierarchical practice culture of openness, in which individuals are willing to challenge (possibly senior) colleagues’ opinions, and to be challenged. Training needs also need to be addressed.^10 12^ * However, clinicians learn from reflecting on and discussing cases (good practice as well as mistakes) **(mechanism)**, and interdisciplinary knowledge-sharing might help identify the role of practice team members in promoting early diagnosis.^5 10^ **(outcome)**  Within this type of practice culture, GPs appear to gain confidence from mutual support – they feel reassured by knowing that a colleague would have taken the same action. Other clinical and non-clinical staff also feel supported and are encouraged to report their suspicions to the GP, including those based on ‘intuition’. For example, it is recognised that long-serving reception staff know the patients very well and this experience is valued. However, training in navigation and red flags would be an advantage.* | Where usual practice includes discussion of patient-safety (e.g. safety ‘huddles’, audit, SEA, reflection), and there is a culture of frank, informal, non-judgemental interaction **(context)**, and by sharing experiences and supporting each other, GPs benefit in terms of improved knowledge and confidence in their actions **(mechanism)** then they will be more likely to recognise when investigations and referrals are warranted, and less likely to delay referral because of uncertainty **(outcome)**. |
| **Conjectured CMO 5** | **Evidence** | **Refined CMO 5** |
| Where a practice audit of cancer referrals is conducted and feedback is given to GPs on whether recommended investigations and examinations were conducted **(context)**, they will be aware of and reflect on their own personal performance, compared with local or national performance, and recognise how it can be improved and **(mechanism)** then they will be more likely to perform these tasks consistently, in line with good clinical practice and NICE guidance **(outcome)**. | Evidence suggests that a focussed examination should be a fundamental component of every consultation unless there is justification to omit it, however examinations were often omitted.^6^ Discussion with patients was mentioned as an important factor in deciding whether they need investigations or not; practices where doctors were rated highly for communication skills tended to use investigations more frequently.^13^* GPs’ knowledge of the guidelines is imperfect, partly due to the large volume of guidance. Furthermore, some GPs consider clinical experience/judgement to be more important than following guidelines. Time constraints and the imperative to refer, and not waste time, also mitigate against investigations.^2 10 12^* **(context)** *The evidence in this theory area is focussed on the role/usefulness of tests and examinations, rather than audit of them, therefore there is no evidence (so far) where audit is the trigger for the awareness of and improvement in performing the recommended examinations and tests.* **(mechanism)** For cancers that have few or vague symptoms in the early stages, simple tests and examinations, which are easy and cheap, can reveal significant findings.^5^ **(outcome)**  In order to be effective, it is important that thorough examination is supported by good history taking and listening to the patient. Knowledge of the guidelines may be improved by CME. Summary booklets may also be helpful but there is a mixed response to electronic reminders, with some GPs finding them annoying. Some GPs were uncertain of their skills in relation to some physical examinations (abdominal, PR, use of dermatoscope and fundoscopy were mentioned), and about when to reinvestigate when tests come back negative but symptoms don’t resolve. There were concerns about over-investigating as well as under-investigating, some GPs were inclined to do many in order to strengthen a referral and risk having it down-graded, but this could delay the referral.* | Where GPs’ educational and skills needs are addressed regarding the examinations and tests advised by NICE in relation to suspected cancer, they will be aware of the recommended investigations and how to conduct them (though guidance is complex and subject to change, so additional summary notes and reminders may be appropriate) **(context)**. Then they will be able to employ the relevant knowledge and skills in their assessment of patients **(mechanism)** and, in the absence of reasons not to investigate, they may choose to do so more in line with NICE guidance (**outcome)**. |
| **Conjectured CMO 6** | **Evidence** | **Refined CMO 6** |
| In the face of uncertainty about individual cancer referral decisions (e.g. vague symptoms, low-risk but not no-risk), if a philosophy of cancer-risk-averseness, and a practice culture of case-finding rather than gate-keeping are engendered **(context)**, GPs will be more likely to lower their individual risk threshold for urgent referrals and Think Cancer! **(mechanism)**, so that more cases of suspected cancer will be identified and referred **(outcome)**. | Evidence supports the consideration of referral for any patient with suspicious symptoms, e.g. unexplained weight loss, new and persistent back pain, or anaemia, even if initial investigations appear normal.^2 12^ **(context)** The usual diagnostic process (i.e. diagnosis based on what’s most likely) works well if the diagnosis isn’t held too rigidly.^1 9 14 15^ **(mechanism)** If the threshold for considering referral is lowered then, when the diagnosis is uncertain, cancer is not ruled out and, when the patient is at increased risk (for reasons of age, co-morbidity or other factors), the GP is more cautious.^1 12^ **(outcome)**  Keeping an open mind (not ruling cancer out too early), and having a practice culture that supports a case-finding approach (as opposed to gate-keeping) are crucial. Unfortunately, the gate-keeping culture is fairly well ingrained in general practice.^11^* Good communication and history taking help: practices where doctors were rated highly for communication skills tended to use urgent cancer referrals more frequently; both GPs and patients tend to normalise symptoms; some patients exaggerate or underplay their symptoms, some don’t want to bother the doctor.^13^* However, the rarity of some cancers, and commonness of associated symptoms makes these cancer harder to suspect,^4 10^* and false-negative test results may further obscure the picture.^8 12^ Furthermore, rigid referral criteria and lack of understanding among policy makers and those implementing cancer initiatives about how cancer risk is managed in primary care (e.g. leading to down-grading of referrals) hamper early referral.^6 11^ * The value of ‘gut feeling’ in the recognition of early cancer is recognised,^6 11^* but not necessarily appreciated in secondary care. | In the face of uncertainty about individual cancer referral decisions (e.g. vague symptoms, low-risk but not no-risk), if a philosophy of cancer-risk-averseness, and a practice culture of case-finding rather than gate-keeping are engendered **(context)**, GPs will be more likely to lower their individual risk threshold for urgent referrals and Think Cancer! **(mechanism)**, so that more cases of suspected cancer will be identified and referred earlier **(outcome)**. |

* Evidence supported by GP interviews, practice focus groups and/or free-text comments from survey.

13. Lyratzopoulos G, Mendonca SC, Gildea C, McPhail S, Peake MD, Rubin G, et al. Associations between diagnostic activity and measures of patient experience in primary care: a cross-sectional ecological study of English general practices. *Br J Gen Pract*

2017;https://doi.org/10.3399/bjgp17X694097.

14. Davies M. Five things I wish I’d known at the start of my career as a GP. 2017:BMJ 2017;357:j3042 doi: 10.1136/bmj.j3042.

15. Sirota M, Kostopoulou O, Round T, Samaranayaka S. Prevalence and alternative explanations influence cancer diagnosis: An experimental study with physicians. *Health Psychology* 2017;36(5):477-85.
