## Supplementary File 3 for "ThinkCancer! The multi-method development of a complex behaviour change intervention to improve the early diagnosis of cancer in primary care"

Supplementary File 3 Behaviour Change Wheel Method and Results

Figure 1 illustrates the different layers that form the Behaviour Change Wheel, with the COM-B model at its centre. COM-B describes how changing behaviour at the individual and/or system level is a result of changing one or more components of psychological and physical Capability, social and physical Opportunity, and automatic and reflective Motivation. Detailed methodology of the application of the behaviour change wheel in the WICKED programme intervention development process and results is at (Supplementary File 3)The COM-B Model is encircled and supported by the Theoretical Domains Framework (TDF) layer (represented in yellow), which provides a deeper level of understanding of the factors (or sources) that influence behaviour, and consists of fifteen theory domains that dovetail with the three COM-B components. Together, the COM-B model and TDF are used to identify what needs to change in order for the target behaviour to be achieved. The next layer (in red) describes nine potential functions that the intervention/s could serve, with each COM-B component and TDF domain linked to an intervention function. The outermost layer of the Behaviour Change Wheel (shown in grey) describes seven policy categories, or actions taken by responsible authorities, that could enable or support the intervention. At each layer of the Behaviour Change Wheel, complementary tools (e.g. Intervention Functions Matrix, APEASE criteria, Behaviour Change Techniques Taxonomy) are available to guide developers’ decisions about intervention functions, specific content and delivery.

Figure 1: The Behaviour Change Wheel

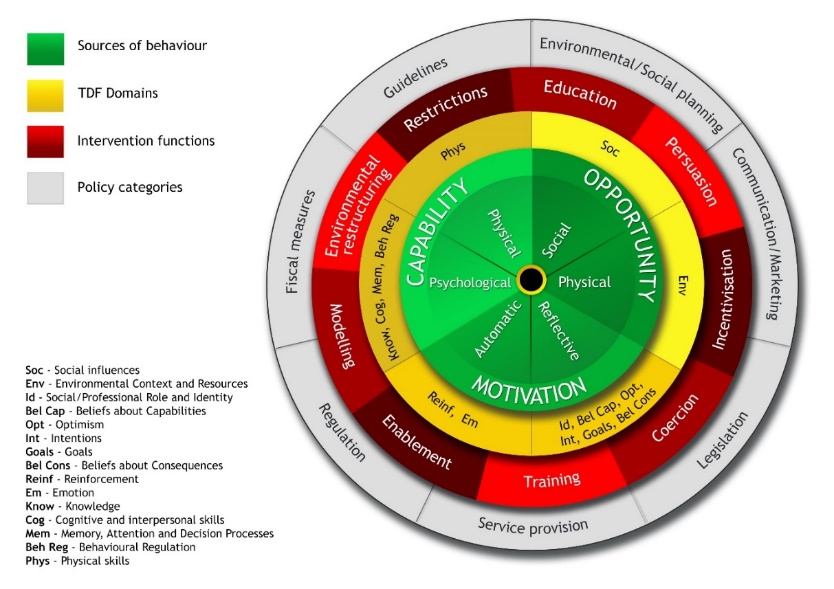

The overall process of applying the Behaviour Change Wheel involved a core working group, with ‘data owners’ (i.e. the researchers who had collected the data for specific elements) allocated to each source of WP1 and WP2 evidence. Tasks such as mapping the sources of behaviour to COM-B and TDF were initially completed independently by at least two individuals in order to reduce subjectivity, followed by core group discussion to arrive at a consensus. Different parts of the Behaviour Change Wheel were applied in stages. Firstly both Work Package 1 and Work Package 2 evidence was reviewed as follows:

- The matrix of evidence synthesised in the realist review
- Key findings from the quantitative survey, including free-text comments
- Coded data from the qualitative interviews and focus groups

Three data-owners scrutinised the data sets to identify all possible influences (barriers and facilitators) upon the target behaviour. The influences were listed, to populate a mapping spreadsheet, with brief explanatory notes to aid understanding. Subsequently, they were mapped, independently by three researchers, onto the COM-B components and TDF domains to identify which intervention functions are likely to be most effective in changing the target behaviour. The spreadsheet was then reviewed in two rounds of discussion, by two out of three researchers, in order to:

- Discuss differences and agree the mapping decisions
- Identify duplication or overlap in the influences and remove or combine as appropriate
- Identify and remove influences that were considered not to be modifiable by the planned ThinkCancer! intervention

The non-modifiable Influences were collated in a supplementary file and categorised by theme. The original list of 150 influences was reduced to 33.

Using the matrix of intervention functions, informed by the definitions of intervention functions ^39^ we were then able to identify the relevant intervention functions that corresponded to the COM-B components identified at the mapping stage. We then applied the APEASE criteria (Affordability, Practicability, Effectiveness/cost-effectiveness, Acceptability, Side effects/safety and Equity) to each of the intervention functions, once again independently by two researchers, followed by discussion and agreement. The selected intervention functions were **education, training, enablement, environmental restructuring, modelling, persuasion and incentivisation.**

Intervention functions were then matched to relevant Behaviour Change Techniques (BCTs), for example, in relation to education and training, the relevant intervention functions included: *instruction, information about health consequences, feedback on outcomes, prompts/cues, self-monitoring, and verbal persuasion about capability*. The same two researchers applied the APEASE criteria to the selected BCTs as before. Subsequently, two researchers considered the intervention functions and behaviour change techniques that met the APEASE criteria alongside the influences on the target behaviour for each TDF/COM-B category. Thus a set of initial ideas for intervention type (from the intervention function exercise) and specific intervention content (from the BCTs) were proposed. These ideas were incorporated in a simplified, one-page table, used as a discussion document for a meeting of an intervention development group comprising the core operational group plus GP stakeholders and patient/public representative. The intervention elements described were brought together in an intervention visualisation (see below). Further development and refinement of the intervention was discussed within subsequent meetings of the intervention development group and actioned by the operational group.

Proposed intervention components and supporting BCT

| **Possible intervention component** | **Behaviour change techniques** | **Application** |
| --- | --- | --- |
| Workshop | Information about health consequences  Problem solving  Action planning  Could be multiple other BCTs here, depending on content/aims of workshop | Increase knowledge and awareness re importance of early diagnosis.  Develop a practice-specific safety-netting plan (could be core components + selected ones). |
| Educational sessions | Information about health consequences  Credible source  Verbal persuasion about capability | Increase knowledge and awareness.  Cancer presentation, early presentations in particular and red flag symptoms and signs.  Dangers and appropriate management of false negative test results.  Support for GPs’ confidence in referral.  Could be tailored according to individual and/or practice needs. |
| Practical instruction | Instruction on how to perform a behaviour | How to operationalise the safety-netting plan. (Part of workshop?)  Address lack of confidence in examination skills. (?APEASE) |
| Online teaching resources | Information about health consequences  Credible source | Could tie in with educational sessions above. Could be signposting to existing resources rather than provision of new. |
| Self-assessment exercise | Feedback on outcome(s) of the behaviour  Prompts/cues  Self-monitoring of behaviour | Focussing on cancer presentation, early presentations in particular and red flag symptoms and signs. |
| Sharing learning from significant event analysis and/or dissemination of examples via cluster groups, as email update, eBulletin or newsletter | Feedback on outcome(s) of the behaviour  Restructuring of the physical environment  Problem solving | Fostering a culture of shared knowledge/learning/experience, openness and frank discussion etc. |
| Whole practice team approach:  “Cancer aware” whole practice team training and award i.e. “we are a cancer aware practice”.  Appointment of role model in “cancer champion” at practice and/or cluster level | Verbal persuasion about capability  Restructuring the social environment  Information about health consequences  Prompts/cues  Action planning | Awareness-raising, confidence-building. Promoting team spirit, collective diligence etc.  Promote safety-netting culture and practice.  Confidence building in use of language and attitude towards the word “cancer” and empowerment of all staff to share concerns  Common goal and ethos for practice  Might need organisational changes to support new roles/ responsibilities. |
| Safety-netting systems/processes e.g. use of safety-netting ‘prescription’. | Restructuring the physical environment  Adding objects to the environment | Allow patient and practice staff to be more vigilant and accepting of early clinical review. Could be , tailored by selection from a ‘toolkit’ of possible elements. |
