## Supplementary File 4 for "ThinkCancer! The multi-method development of a complex behaviour change intervention to improve the early diagnosis of cancer in primary care"

Supplementary File 4 – Review of reviews - characteristics of the included studies and summary of findings

| **Author, year**  **country** | **Review aims (including cancer type)** | **Methods** | **Author's conclusion / Relevant findings** |
| --- | --- | --- | --- |
| Abel, 2008  USA | To describe a possible problem regarding referral and diagnosis of patients with **haematological malignancies** by comparing with solid tumours (**breast and colon**). | Systematic review  Clearly framed question: Yes  Number of databases searched: 3  Year of searches: Not reported  Search strategy or search terms provided: Yes  Duplicate process: Not reported  Study selection criteria described: No  Types of studies: CCtr; CCoh; HCoh  Data synthesis: Narrative | No of included studies: 56  Quality appraisal: not reported  Lack of primary care professional exposure to, and knowledge of, the chronic hematologic malignancies as well as possible biases due to patient characteristics (such as gender, extremes of age, and race-ethnicity) all likely play a role in delayed diagnosis. Patients who present with symptoms are more likely to be referred than those who are asymptomatic, and patient delay in seeking care also seems important. |
| Brown, 2014  UK | To compare the characteristics of **healthcare systems** in International Cancer Benchmarking Partnership (ICBP) jurisdictions, as they relate to cancer diagnosis, to identify characteristics that would plausibly modify the diagnostic pathway, and thereby outcomes, for patients with suspected cancer. | Consensus process with key informants (ICBP Board members for each jurisdiction) based on an earlier literature review, the methods of which are not reported or referenced. | Differences in healthcare systems (such as regulation, financing, the gatekeeping role of GPs, direct access to secondary care, the degree of comprehensiveness of primary care services provided, the level of cost sharing, and the type of primary care providers) are not systematically associated with differences in cancer outcome. Factors that could have an influence on patient and professional behaviour, and consequently contribute to delays in cancer diagnosis and poorer cancer outcomes in some countries, include centralisation of services, free movement of patients between primary care providers, access to secondary care, and the existence of patient list systems. |
| Burge, 2016  UK | To identify areas of clinical practice where change may influence **bladder cancer** outcomes. | Systematic review  Clearly framed question: Yes  Number of databases searched: 1  Year of searches-inception–Dec 2015  Search strategy or search terms provided: Yes  Duplicate process: Not reported  Study selection criteria described: Yes  Types of studies: not reported  Data synthesis: Narrative | No of included studies: 55  Quality appraisal: not reported  The review identified evidence of poor recognition of the signs and symptoms of bladder cancer, a difference in referral patterns between genders and a possible difference in treatment stratagems. Most studies indicate that men are referred more promptly; women have more pre-referral consultations. The presence of cystitis, in particular, delays referral. |
| Carter, 1998  UK | To explore the reasons behind the delay in presentation of **colorectal carcinoma**. | Traditional literature review | Reasons for longer times to diagnosis in primary care included examinations not being performed because of a reluctance on the part of the patient, expectation of a repeat rectal examination at the hospital, insufficient time, or lack of a chaperone. In one study, delay in referring a patient was increased if the patient had only one symptom, had diarrhoea, or was of low social class, whereas if the patient had constipation or was middle class delay was less likely. |
| Gikas, 2014  Greece | To explore the existing and potential roles of the primary care professional in the diagnosis and management of GI disorders (Not specific to cancer). | Traditional literature review | The review identified evidence that primary care professionals don't always investigate upper GI symptoms, e.g. ‘In a study in the UK, it was noticed that 47% of 431 patients presenting to their general practitioner with an iron-deficient anaemia were adequately managed’. The authors advise that the identification of high-risk patients and prompt endoscopic examination are the best strategies for early diagnosis of upper GI malignancies, and suggest that an accurate and complete medical history (including family history) in combination with detailed physical examination remains the most critical aspect of diagnosing GI malignancies. They conclude that updating of knowledge and skills of primary care professionals via continuing medical education is the only way for better adherence with standards and improving quality of care for patients with gastrointestinal diseases. |
| Hamilton, 2010 UK | To examine expedited diagnosis of symptomatic cancer from four perspectives. 1) The potential for clinical benefits. 2) How it can be achieved. 3) The most appropriate patients for cancer investigations, and the possible community settings for identification of such patients. 4) The health economics. | Traditional literature review | The authors note that some cancers, e.g. myeloma, are notoriously difficult to diagnose and there are often multiple consultations in primary care. Counterintuitively, continuity of primary care has only a very small effect upon the rapidity of a cancer diagnosis. Clinical decision support (using risk-assessment tools) improves physician performance and the ordering of diagnostic tests. Such tools can, however, conflict with clinical judgement, making some GPs reluctant to use them in the consultation, and are trusted less by more experienced GPs. Furthermore, variation in interpretation of symptoms by different clinicians can lead to substantial variations in risk assessment. The authors conclude that some of the improvements in cancer survival are almost certainly a result of improved diagnostics, major reconfigurations and investment in cancer services, and a liberalisation of the criteria for cancer investigation, coupled with better identification of the individuals who are most at risk. |
| Macdonald, 2006 UK | To evaluate factors associated with the time interval between patient and primary care practitioner delay for **upper gastrointestinal cancer**; or to describe an intervention designed to reduce those intervals. | Systematic review  Clearly framed question: Yes  Number of databases searched: 7 + grey literature  Year of searches-1970 to Nov 2003  Search strategy or search terms provided: Yes  Duplicate process: Yes  Study selection criteria described: Yes  Types of studies: CCtr; CCoh; HCoh; Qual  Data synthesis: Narrative | No of included studies: 25 (5 studies with multiple cancers)  Quality appraisal: 6 strong, 8 moderate, 5 poor, based on authors’ assessment of methods  Studies reporting delay intervals demonstrated that the patient phase of delay was greater than the practitioner phase, whilst patient-related research suggests that recognition of symptom seriousness is more important than recognition of the presence of the symptom. The main factors related to practitioner delay were misdiagnosis, application and interpretation of tests, and the confounding effect of existing disease. No relevant intervention studies were identified. |
| Mangion, 2014  UK | To explore the relationship between the ‘two week rule’ (TWR) and the diagnosis of **colorectal cancer** (CRC). | Systematic review  Clearly framed question: Yes  Number of databases searched: 3  Year of searches-1999-2013  Search strategy or search terms provided: Yes  Duplicate process: not reported  Study selection criteria described: Yes or No  Data synthesis: Narrative | No of included studies: 12  Quality appraisal: not reported  The authors found that delay by GPs is often due to their failure to recognise CRC signs and symptoms, and there is a subsequent delay in referring these patients to appropriate clinics. An objective of the TWR was to detect CRC at an earlier disease stage, but the literature was inconclusive as to whether or not this had been achieved. reasons for these findings include the lack of highly specific clinical guidelines, which may be misinterpreted by GPs. Lack of GP awareness of the TWR system had also been implicated as a cause of the under-use of the TWR referral route, |
| Mansell, 2011  UK | To identify interventions that reduce primary care delay in the referral of patients with cancer to secondary care. (**any cancer**) | Systematic review  Clearly framed question: Yes  Number of databases searched: 8  Year of searches: inception – Mar 2010  Search strategy or search terms provided: Yes  Duplicate process: Yes  Study selection criteria described: Yes  Types of studies: RCT; q-RCT; non-RCT; CCtr; CCoh; HCoh  Data synthesis: Narrative | No of included studies: 22  Quality appraisal: 6 good, 12 moderate, 4 poor, based on Newcastle-Ottawa Quality Assessment tool.  This review covered any cancer, though only skin, breast, colorectal and prostate studies were found. Interventions included in the narrative synthesis were education, audit and feedback, decision support software and guideline use, and other training. The authors identified 22 interventions but there was no evidence that any intervention directly reduced primary care delay in the diagnosis of cancer. Limited evidence suggests that complex interventions, including audit and feedback and specific skills training, have the potential to do so. |
| Mitchell, 2008  UK | To explore factors influencing pre-hospital delay – the time between a patient first noticing a cancer symptom and presenting to primary care or between first presentation and referral to secondary care. (**any cancer**) | Systematic review  Clearly framed question: Yes  Number of databases searched: 9 + grey literature  Year of searches: 1970 - 2003  Search strategy or search terms provided: Yes  Duplicate process: Yes  Study selection criteria described: Yes  Types of studies: CCtr; CCoh; HCoh; Qual  Data synthesis: Narrative | No of included studies: 54 (12 with multiple cancers)  Quality appraisal: 24 strong, 15 moderate, 4 poor; appraisal tool not reported  Initial misdiagnosis, inadequate examination and inaccurate investigations increased practitioner delay. Use of referral guidelines may reduce delay, although evidence is currently limited. No intervention studies were identified. The authors concluded that, if delayed diagnosis is to be reduced, there must be increased recognition of the significance of symptoms among patients, and development and evaluation of interventions that are designed to ensure appropriate diagnosis and examination by practitioners. |
| Mitchell, 2015  UK | To identify patient and practitioner factors that influence **colorectal and lung** cancer diagnosis via emergency presentation (EP). | Systematic review  Clearly framed question: Yes  Number of databases searched: 8  Year of searches: 1996 - Mar 2014  Search strategy or search terms provided: Yes  Duplicate process: Yes  Study selection criteria described: Yes  Types of studies: CCtr; CCoh; HCoh  Data synthesis: Narrative | No of included studies: 22  Quality appraisal: 15 Strong, 3 moderate, 4 poor, based on Newcastle-Ottawa Quality Assessment tool.  The authors conclude that certain patient-related factors, such as age, gender and socioeconomic deprivation, have an influence on diagnosis of cancer during an EP. It also shows that cancer symptoms and patterns of healthcare utilisation are relevant. However the context in which risk factors for EP exist remain unclear and need to be better understood. |
| Ott, 2009  Canada | To assess the consistency and availability of a definition for early cancer symptoms as well as to assess the impact of early cancer diagnosis on survival. (**any cancer**) | Traditional literature review | The authors present a detailed list for several cancers of symptoms associated with early diagnosis, and those associated with later stage. They conclude that addressing the reasons for late cancer diagnosis is a particular challenge for primary care, which is usually the patient’s first contact with the health system and the area to which the delay is applicable. This also involves significant challenges for practitioners in assessing cancer symptoms to avoid misdiagnoses that contribute to delays. |
| Schichtel, 2013  UK | To examine the evidence of effectiveness of educational interventions for primary healthcare professionals to promote the early diagnosis of cancer. (**any cancer**) | Systematic review  Overall quality: High  Clearly framed question: Yes  Number of databases searched: 9 + grey lit  Year of searches-1948–Apr2012  Search strategy or search terms provided: Yes  Duplicate process: Yes  Study selection criteria described: Yes  Types of studies: RCTs  Data synthesis: Narrative | No of included studies: 20  Quality appraisal: 5 good, 13 moderate, 2 poor; appraisal tool not reported.  The authors found some evidence to inform the development of further educational interventions to promote the early diagnosis of cancer in primary care. RCTs including interactive education, reminder systems and audit and feedback had positive effects. They discovered that RCTs including interactive education, reminder systems and audit and feedback had positive effects. However only short-term effects were shown. Long-term effects, and effectiveness in any cancer (as opposed to specific cancers) remain unclear. |
| Schmidt-Hansen, 2015  UK | To quantify the risk of urinary tract cancer in patients presenting in primary care with symptoms that may indicate **bladder or renal cancer**. | Systematic review  Clearly framed question: Yes  Number of databases searched: 6  Year of searches-1980 to Aug, 2014  Search strategy or search terms provided: Yes  Duplicate process: Yes  Study selection criteria described: Yes  Types of studies: CCtr; CCoh; HCoh  Data synthesis: Narrative | No of included studies: 11  Quality appraisal: 5 Low risk of bias, 6 High, based on Cochrane tool  Review presented relevant symptoms in primary for **bladder and renal tract cancers**. Apart from haematuria (which tends to be investigated anyway), none of the symptoms had a high positive predictive value. The review was undertaken as part of the process to revise UK guidance. |
| Shapley, 2010  UK | To identify symptoms, signs, and non-diagnostic test results in unselected primary care populations that are highly predictive of cancer. (**any cancer**) | Systematic review  Overall quality: High  Clearly framed question: Yes  Number of databases searched: 14 + hand searches  Year of searches: inception - Oct 2009  Search strategy or search terms provided: Yes  Duplicate process: Yes  Study selection criteria described: Yes  Types of studies: CCtr; CCoh; HCoh  Data synthesis: Meta-analysis and Narrative | No of included studies: 25  Quality appraisal: 19 good, 6 poor, based on Newcastle-Ottawa Quality Assessment tool.  There is robust evidence for eight symptoms, signs, and non-diagnostic test results as strongly indicative of cancer for specific age and sex groups in unselected primary care populations. These have the potential to improve the early diagnosis of some cancers in primary care by the use of computer warning flags, improved guidelines, audit, and appraisal. |
| Thompson, 2010 UK | To determine current delays in diagnosis and treatment of **bowel cancer**, when and why they occur, and what effect they have on survival | Systematic review  Clearly framed question: Yes  Number of databases searched: 6 + hand searches  Year of searches: 1990-2008  Search strategy or search terms provided: Yes  Reference management described: Yes  Duplicate process: Yes  Study selection criteria described: Yes  Types of studies: not reported  Data synthesis: Narrative | No of included studies: 80  Quality appraisal: 0 grade 1, 1 grade 2, 12 grade 3, 8 grade 5, 18 grade 5, based on criteria developed from Grade recommendations for other conditions, where grade 1 is good and grade 5 is poor.  Review concludes that campaigns to earlier diagnose bowel cancer will not be successful unless new strategies are developed. There is substantial evidence that earlier diagnosis of symptomatic bowel cancer will not improve survival in the majority of patients. However as excessive delays still occur in some patients it is reasonable to continue to aim to diagnose and treat all bowel cancer within 6 months of the onset of symptoms with an overall median of 3–4 months.  The paper present a moderately strong argument that early diagnosis of symptomatic colorectal cancer (and perhaps endometrial, melanoma, cervical, and breast) is futile, as the cancer's biological nature determines outcome before presentation with symptoms |
| Williams, 2016  UK | To systematically identify and compare the performance of models that predict the risk of primary **colorectal cancer** (CRC) among symptomatic individuals. | Systematic review  Clearly framed question: Yes  Number of databases searched: 2 + hand searches  Year of searches-Jan 2000-Mar 2014  Search strategy or search terms provided: Yes  Reference management described: Yes  Duplicate process: Yes  Study selection criteria described: Yes  Types of studies: CCtr; CCoh; HCoh  Data synthesis: Narrative | No of included studies: 18  Quality appraisal: 11 High, 4 moderate, 3 Low, based on Critical Appraisal Skills Programme (CASP) tool.  Models with good discrimination have been developed in both primary and secondary care populations. Most contain variables that are easily obtainable in a single consultation, but further research is needed to assess clinical utility before they are incorporated into practice. |

Types of studies: Randomised Controlled Trial (RCT); Quasi-Randomized Controlled Trial (q-RCT); Non-Randomised Controlled Trial (non-RCT); Case Control (CCtr); Current Cohort (CCoh); Historical Cohort (HCoh)
