## Supplementary File 5 for "ThinkCancer! The multi-method development of a complex behaviour change intervention to improve the early diagnosis of cancer in primary care"

Supplementary File 5 Discrete Choice Experiment

Table 1: Attributes and attribute levels used in the study

| **Attributes** | **Levels** | **Definition and effects coding ( )** |
| --- | --- | --- |
| Type of prompt | Non-computerised prompt | Non-computerised prompt e.g. mouse mat, info sheet or poster describing symptoms or detailing risk score; printed NICE summary guidelines /Macmillan materials. (base) |
|  | Computerised prompt | Computerised prompt that flags risk scores (e.g. risk prediction models such as ‘Q-Cancer’ or Macmillan prompts) in medical records which are informed by NICE guidelines or when abnormal symptom(s) identified. |
| Type of audit | Practice level | Practice level e.g. individual practice. (base) |
|  | Cluster level | Cluster level e.g. a number of geographically clustered practices |
|  | National level | National level e.g. NHS Wales |
| Type of education and training | covering recognition of signs and symptoms | Education and training covering recognition of signs and symptoms and spotting them. (base) |
|  | covering the use of diagnostic tests | Education and training covering the use of diagnostic tests and how to interpret their results. |
| Mode of education and training delivery | Internet-based | Internet-based learning such as online self-learning. (base) |
|  | Face-to-face | Face-to-face learning such as practice-based learning, attending workshops, conferences, lectures, etc. |
|  | Both | Both face-to-face and internet-based learning e.g. online self-learning. |
| Access to diagnostic testing for GPs | Access predominantly through secondary care | Access to diagnostic testing predominantly through secondary care. (base) |
|  | GPs have open access | GPs have open access to diagnostic tests and responsibility for interpretation of results and subsequent actions. |
| Safety netting | Patient responsibility | Patient responsibility for subsequent behaviour and actions (e.g. following up blood results and making further appointment if no better or symptoms persisting). (base) |
|  | Primary care responsibility | Primary care responsibility for following up blood results and/or checking to see if patient getting better or not, including use of IT, and checking on referral sent and investigations / appointments attended. |
| Average amount of time able to spend, in an ideal world, on activities to achieve timely cancer diagnosis | 0.5 hour per week | 0.5 hour per week on average in an ideal world. (30) |
|  | 1 hour per week | 1 hour per week on average in an ideal world. (60) |

Table 2 Discrete Choice Experiment (DCE) - results from the logit regression model: The preferences of GPs for different attributes (characteristics) surrounding timely diagnosis of cancer in primary care: WICKED study (n = 151)

|  | **WICKED DCE study sample (n = 151)** | | |
| --- | --- | --- | --- |
| **Attribute^§^** | **β-coefficient** | **95% CI^¥^** | **P-value** |
| Access to diagnostic testing for GPs – Access predominantly through secondary care (omitted category)**^∆^** | -0.2218***** | -0.2866 to -0.1930 | 0.000 |
| **Access to diagnostic testing for GPs – GPs have open access^Ω^** | 0.2218***** | 0.1930 to 0.2866 | 0.000 |
| **Type of audit – Practice level^Ω^** (omitted category)**^∆^** | 0.1579***** | 0.0841 to 0.2585 | 0.000 |
| Type of audit – Cluster level (*Regional grouping practices*) | 0.0245 | -0.0481 to 0.1012 | 0.493 |
| Type of audit – National level | -0.1824***** | -0.2699 to -0.1231 | 0.000 |
| Type of prompt – Non-computerised prompt (omitted category)**^∆^** | -0.1206***** | -0.1749 to -0.0857 | 0.000 |
| **Type of prompt – Computerised prompt^Ω^** | 0.1206***** | 0.0857 to 0.1749 | 0.000 |
| Mode of education and training delivery – Internet (omitted category)**^∆^** | -0.1143***** | -0.2072 to -0.0385 | 0.001 |
| Mode of education and training delivery – Face-to-face | -0.0076 | -0.0796 to 0.0674 | 0.832 |
| **Mode of education and training delivery – Both^Ω^** | 0.1219***** | 0.0647 to 0.2013 | 0.001 |
| **Safety netting – Patient responsibility^Ω^** (omitted category)**^∆^** | 0.1043***** | 0.0698 to 0.1560 | 0.000 |
| Safety netting – Primary care responsibility | -0.1043***** | -0.1560 to -0.0698 | 0.000 |
| Type of education and training – Covering recognition of signs and symptoms (omitted category)**^∆^** | 0.0209 | -0.0207 to 0.0693 | 0.337 |
| Type of education and training – Covering the use of diagnostic tests | -0.0209 | -0.0693 to 0.0207 | 0.337 |
| Time able to spend on activities to achieve timely cancer diagnosis | 0.0006 | -0.0022 to 0.0039 | 0.682 |
| Constant | -0.0808 | -0.1906 to 0.0148 | 0.211 |
| Number of obs = 2416 | | | |
| Number of groups = 151 | | | |
| Wald chi2(9) = 180.58 | | | |
| Log likelihood = –1561.91 | | | |

**^∆^** The β-coefficient on the omitted level of an effects-coded variable is calculated as the negative sum of the β-coefficients on the non-omitted levels of that attribute

***** P < 0.05

**^¥^** 95% confidence intervals generated using non-parametric bootstrapping

**^§^** The significant attributes (p<0.05) are placed in order of importance, from most to least important.

**^Ω^** The highlighted attribute level (font in bold) shows the preferred level for that particular significant attribute
