## Supplementary figures and images for "ThinkCancer! The multi-method development of a complex behaviour change intervention to improve the early diagnosis of cancer in primary care"

### Supplementary File 6

Supplementary File 6 CRUK Safety netting flow diagram (Reproduced with permission)

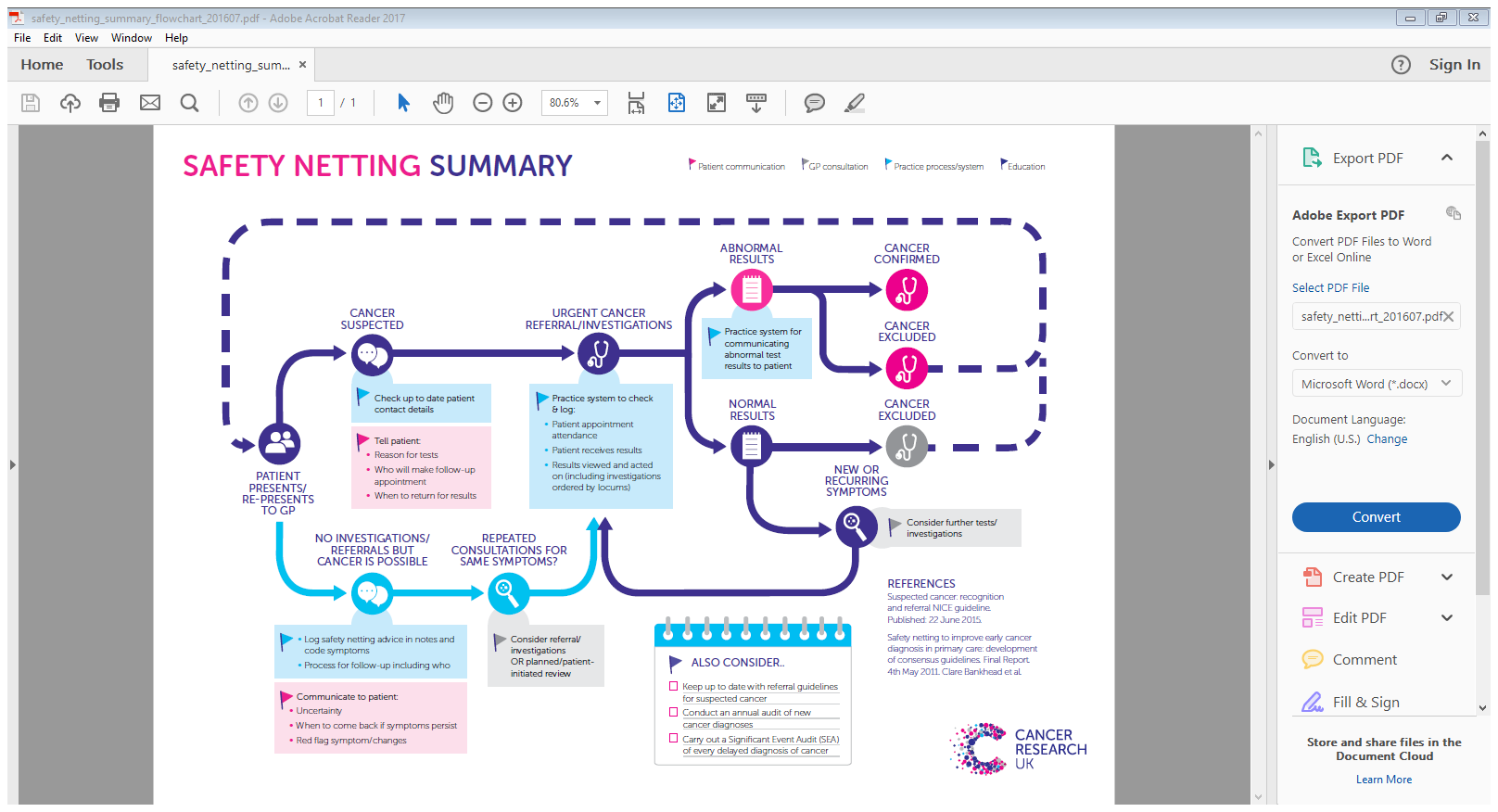
